# Estimated Sp02/Fio2 ratio to predict mortality in patients with suspected COVID-19 in the Emergency Department: a prospective cohort study

**DOI:** 10.1101/2020.05.28.20116194

**Authors:** Johannes Von Vopelius-Feldt, Daniel J Watson, Carla L Swanson-Low, James A Cameron

## Abstract

**Background:** This study examined whether the presence and severity of Type 1 Respiratory Failure (T1RF), as measured by the ratio of pulse oximetry to estimated fraction of inspired oxygen (SpO2/eFiO2 ratio), is a predictor of in-hospital mortality in patients presenting to the ED with suspected COVID-19 infection.

**Methods:** We undertook a prospective observational cohort study of patients admitted to hospital with suspected COVID-19 in a single ED in England. We used univariate and multiple logistic regression to examine whether the presence and severity of T1RF in the ED was independently associated with inhospital mortality.

**Results:** 180 patients with suspected COVID-19 infection met the inclusion criteria for this study, of which 39 (22%) died. Severity of T1RF was associated with increased mortality with odds ratios (OR) and 95% confidence intervals of 1.58 (0.49 – 5.14), 3.60 (1.23 – 10.6) and 18.5 (5.65 – 60.8) for mild, moderate and severe T1RF, respectively. After adjusting for age, gender, pre-existing cardiovascular disease, neutrophil-lymphocyte ration (NLR) and estimated glomerular filtration rate (eGFR), the association remained, with ORs of 0.63 (0.13 – 3.03), 3.95 (0.94 – 16.6) and 45.8 (7.25 – 290). The results were consistent across a number of sensitivity analyses.

**Conclusions:** Severity of T1RF in the ED is an important prognostic factor of mortality in patients admitted with suspected COVID-19 infection. Current prediction models frequently do not include this factor and should be applied with caution. Further large scale research on predictors of mortality in COVID-19 infection should include SpO2/eFiO2 ratios or a similar measure of respiratory dysfunction.

**KEY MESSAGES:** *What is already known on this topic:* A number of studies have identified potential variables to predict mortality in patients with COVID-19 but have mainly focused on laboratory measures. The primary pathophysiology and cause of death seems to be lung injury leading to an acute respiratory distress syndrome (ARDS) like illness, clinically presenting as type 1 respiratory failure (T1RF). Despite this, only a very small number of studies have included measurements of respiratory dysfunction as predictor of mortality.

*What this study adds:* To our knowledge, this is the first study to examine the association between T1RF as measured by the ratio of pulse oximetry (SpO2) and estimated fraction of inspired oxygen (FiO2) and mortality in patients admitted with suspected COVID-19. The presence and severity of T1RF are strongly associated with mortality. At the same time, other factors previously shown to be associated with mortality are potentially less important than currently assumed, once adjusted for the severity of T1RF.

## BACKGROUND

In December 2019, Wuhan, China, experienced an outbreak of coronavirus disease 2019 (COVID-19), caused by the severe acute respiratory syndrome coronavirus 2 (SARS-CoV-2).[1] Since March 2020, COVID-19 has been declared a pandemic and cases of COVID-19 and associated fatalities are increasing rapidly within Europe and worldwide.[1, 2] A small number of retrospective studies, mainly from China, have attempted to analyse predictive factors of mortality in patients admitted with COVID-19.[3, 4] These studies have uncovered a range of blood tests, demographic features, symptoms, and observations which are associated with mortality from COVID-19 but their usefulness in practice is so far unclear.[5] A recent review of currently suggested prediction models for adverse events from COVID-19 concluded that these models were insufficient but that predictors identified so far should be seen as candidate prognostic factors for further large scale research.[5]

The primary pathophysiology and reason for requiring admission to hospital with COVID-19 seems to be lung injury leading to acute respiratory distress syndrome (ARDS) like illness, clinically presenting as type 1 respiratory failure (T1RF).[2, 6] To our knowledge, none of the currently available research on prognostic factors in the Emergency Department (ED) seems to include the presence or severity of ARDS-like illness. This might be due to the formal definition of ARDS requiring arterial blood gas samples in mechanically ventilated patients, which does not apply to most patients with suspected COVID-19 in the ED. We hypothesise that the presence and severity of T1RF in the ED, as a proxy measure for an ARDS-like illness, could be an important predictor of mortality. This study therefore aims to assess whether a pragmatic approximation of T1RF severity, based on the ratio of peripheral pulse oximetry (SpO2) and estimated fraction of inspired oxygen (eFiO2) can predict mortality in patients presenting to ED with suspected COVID-19.

## METHODS

We undertook a prospective analysis of patients presenting to a single ED in England with suspected COVID-19. The ED sees approximately 80,000 patients per year and is situated in an urban/suburban catchment area with a predominantly Caucasian population. Data was obtained from a prospective clinical effectiveness project aimed at optimising patient flow and resource allocation during the COVID-19 pandemic peak. ED clinicians entered data of patients with suspected COVID-19 onto an online database. The project team then followed up these patients and completed datasets with results of further investigations and patient outcomes, including mortality. Data was anonymised prior to transfer to the research team.

### Inclusion/exclusion criteria

Included in the database were adult patients triaged to a COVID-19 assessment area in the ED and where the treating clinician considered COVID-19 to be at least as likely as the most likely alternative diagnosis. Triage to the assessment area was based on any of the following symptoms: shortness of breath, pyrexia higher than 37.8C or a new persistent cough. From this database, we included in our analysis all patients admitted to hospital and where COVID-19 infection was confirmed or highly likely based on the following criteria published by Public Health England[7]

- At least one nasopharyngeal swab with positive polymerase chain reaction (PCR) result for COVID-19
- Negative COVID-19 PCR results but either chest x-ray or computed tomography (CT) scan reported as ‘classic COVID-19’ according to the British Society for Thoracic Imaging (BSTI) guidelines[8]
- Patients admitted via the above pathway with pyrexia of greater than 37.8C, negative COVID-19 PCR results and indeterminate imaging results, for which the discharge letter or death certification states COVID-19 as most likely diagnosis

We excluded patients who were discharged from the ED or patients still in hospital at the time of analysis.

### Variables recorded

Table 1 provides an overview of variables extracted from the database for all included patients.

**Table 1.** Variables collected on initial data upload to Google spreadsheet

| Patient demographics | ED interventions and disposition |
| --- | --- |
| • Gender<br>• Age group<br>• Rockwood Frailty Score<br>• Co-morbidities<br>• Presenting Symptoms | • Amount of oxygen administered<br>• Intravenous fluids<br>• Steroids or nebulisers<br>• Antibiotics<br>• Non-invasive ventilation<br>• Intubation and ventilation |
| ED observations and investigations | Inpatient course |
| • Vital parameters<br>• Chest x-ray findings<br>• Blood test results<br>• COVID-19 PCR results<br>• CT scan results (if applicable) | • Admission to an intensive care unit<br>• Inpatient death<br>• Inpatient PCR results (if applicable)<br>• Inpatient CT scan results (if applicable) |

*ED: Emergency Department, PCR: polymerase chain reaction, CT: computed tomography*

### Determination of SpO2/FiO2 ratio

FiO2 was estimated from the flow rate of oxygen recorded for all non-intubated patients. Based on limited available evidence, this was done in a pragmatic fashion, with an assumed increase of eFiO2 by approximately 0.04 for every litre of oxygen per minute delivered (see Appendix 1). [9, 10] As arterial blood gases were not routinely taken in ED, T1RF was categorised into none, mild, moderate and severe based on the patient’s recorded SpO2/eFiO2 ratio. Based on previous research in ARDS patients, cut-offs for SpO2/eFiO2 ratios were set to 316, 232, and 148 for mild, moderate, and severe T1RF, respectively.[11] These approximately correspond to PaO2/FiO2 ratio cut-offs for the diagnosis of mild, moderate and severe ARDS of 300, 200 and 100, respectively. In addition, we undertook a sensitivity analysis which used a cut off of SpO2/eFiO2 ratio of 190 (approximately PaO2/FiO2 150) to categorise patients into mild/moderate and moderate/severe T1RF.[12]

### Statistical analysis

We used the chi-square test, Kruskal-Wallis test, and univariate logistic regression to examine distribution of predictive factors in patients who survived to hospital discharge or died in hospital. Two multiple logistic regression models were used to determine if the severity of T1RF as defined by the SpO2/eFiO2 ratio was an independent predictor of mortality. Pre-specified factor selection for model 1 was guided by the strength of existing evidence and included age, gender, pre-existing cardiovascular disease, neutrophil-lymphocyte ration (NLR), estimated glomerular filtration rate (eGFR).[3–6] For the additional model 2, we added further potential factors in a forward-stepwise approach, based on the strength of association with the outcome of interest in univariate analysis. Additional factors considered for model 2 included: Rockwood frailty score, further comorbidities (diabetes, cerebrovascular disease, chronic respiratory disease, active cancer, chronic kidney disease, immunosuppression, obesity), symptoms (days since onset, shortness of breath, cough, headache, myalgia, diarrhoea), observations (body temperature, heart rate, mean arterial blood pressure, respiratory rate), and further blood tests (platelet count, c-reactive protein (CRP), alanine aminotransferase (ALT), troponin, lactate). D-Dimer and ferritin were considered but these blood tests were not obtained routinely in ED and therefore only available for a small subset of patients. Factors were included in model 2 if they demonstrated a statistically significant association once added to model 1. Furthermore, we undertook a sensitivity analysis which only included patients with COVID-19 diagnosis confirmed either by PCR testing or imaging (classic COVID-19 findings on chest x-ray or CT scan according to BSTI guidelines). Model accuracy and goodness-of-fit were assessed using pseudo-R values, Hosmer and Lemeshow goodness-of-fit test, the Akaike Information Criterion (AIC), and area under the receiver-operator curve (ROC).[13] We estimated that a sample size of 150 to 200 patients with 30 to 40 deaths would allow us to specify a multiple logistic regression model with up to five variables.

### Ethics and approvals

The clinical effectiveness project was approved by North Bristol NHS Trust’s Patient Safety, Assurance and Audit Service (reference number CE44619). The need for research ethics committee (REC) review was waived by the Health Research Authority (HRA) based on the fact that only anonymised data was processed for a COVID-19 related research project.[14]

### Patient and public involvement

Due to the time-critical nature of the research project, we were unable to involve patients or members of the public in the planning of this study.

## RESULTS

At the time of analysis, 180 patients with suspected COVID-19 infection met the inclusion criteria for this study. Overall, 39 (22%) patients died and 22 (12%) were admitted to ICU. No patient was mechanically ventilated in the ED at the time of data entry. Positive PCR results were obtained in 117 (65%) of patients, in keeping with previously reported sensitivities for nasopharyngeal PCR swabs.[15] Mortality rates were 10%, 16%, 30%, and 68% in patients with no, mild, moderate, and severe T1RF, respectively. Table 2 confirms that all factors selected a priori to be included in model 1 as potential predictors of in-hospital mortality were indeed unequally distributed between survivors and non-survivors of patients admitted with suspected COVID-19.

**Table 2.**
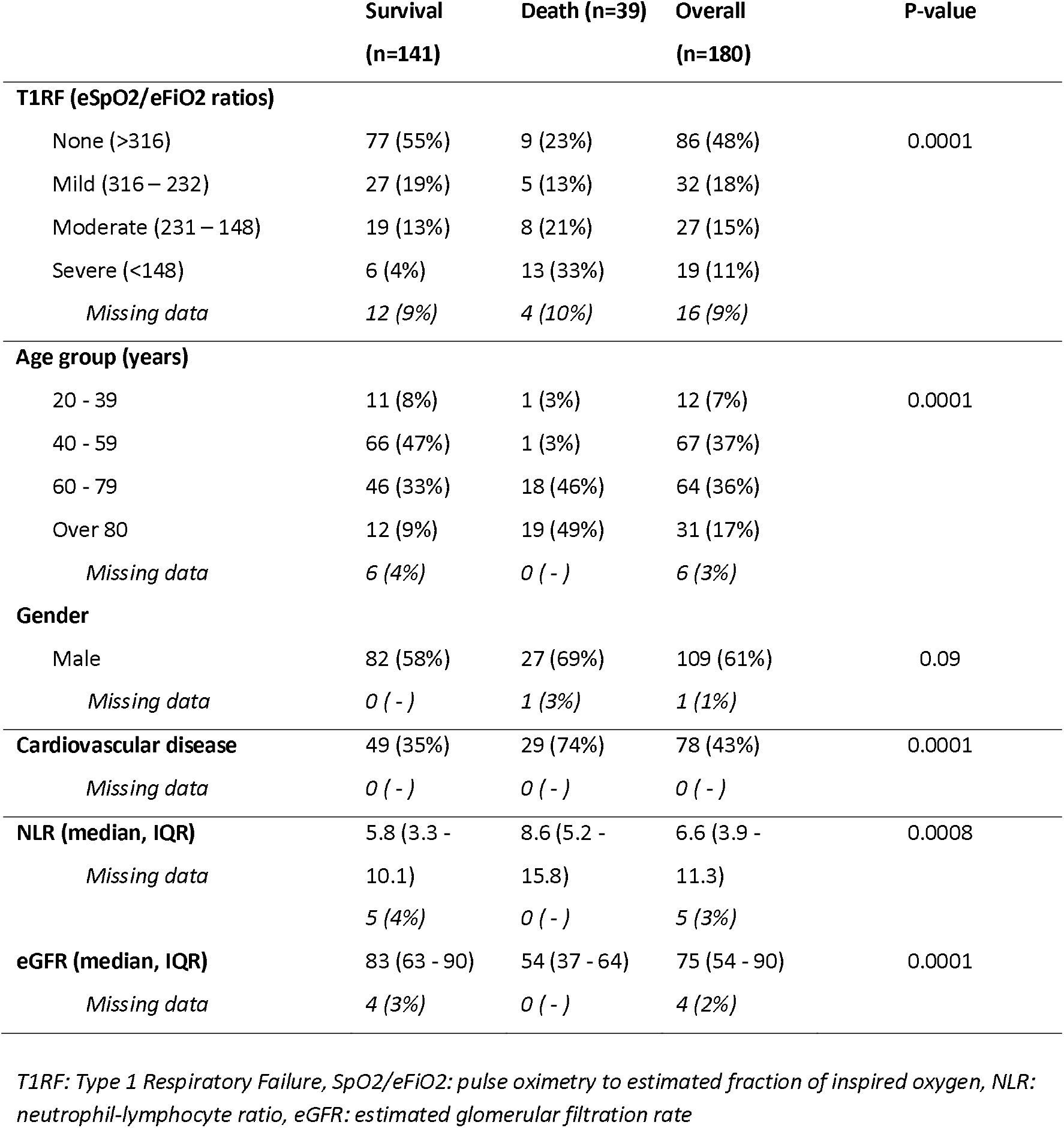
Distribution of previously described predictors of mortality in COVID-19 amongst survivors and non-survivors

Table 3 shows the results of univariate and multiple logistic regression of these factors. In addition, we evaluated further potential factors (see Appendix 2) for inclusion in model 2. Further factors included in model 2 were the presence of immunosuppression (either from medication or disease), body temperature and lactate levels. Both models showed good diagnostic criteria, with model 2 slightly improved over model 1 (see Table 3).

**Table 3.** Association of potential prognostic factors with mortality in patients admitted to hospital with suspected COVID-19.

|  | Univariate analysis | Model 1 (n=158) | Model 2 (n=148) |
| --- | --- | --- | --- |
| **T1RF (SpO2/eFiO2 ratios)** |  |  |  |
| None (>316) | 1 (*reference*) | 1 (*reference*) | 1 (*reference*) |
| Mild (316 – 232) | 1.58 (0.49 – 5.14) | 0.63 (0.13 – 3.03) | 0.18 (0.02 – 1.64) |
| Moderate (231 – 148) | 3.60 (1.23 – 10.6)* | 3.95 (0.94 – 16.6) | 2.42 (0.40 – 14.5) |
| Severe (<148) | 18.5 (5.65 – 60.8)* | 45.8 (7.25 – 290)* | 21.6 (2.34 – 199)* |
| **Age**<br>(per year increase) | 2.48 (1.78 – 3.46)* | 2.45 (1.48 – 4.03)* | 3.16 (1.69 – 5.94)* |
| **Gender**<br>(male) | 1.77 (0.81 – 3.84) | 1.27 (0.39 – 4.15) | 1.96 (0.43 – 8.95) |
| **Cardiovascular disease** | 5.44 (2.45 – 12.1)* | 2.13 (0.61 – 7.41) | 2.02 (0.45 – 9.03) |
| **Immunosuppression** | 3.83 (0.74 – 19.8) | - | 1.95 (0.00 – 8663) |
| **Body temperature**<br>(per degree Celsius increase) | 0.86 (0.67 – 1.12) | - | 0.71 (0.37 – 1.36) |
| **NLR** (per unit increase) | 1.04 (1.01 – 1.08)* | 1.00 (0.94 – 1.06) | 0.98 (0.90 – 1.06) |
| **eGFR** (per unit increase) | 0.96 (0.94 – 0.98)* | 0.97 (0.94 – 1.00)* | 0.97 (0.93 – 1.00)* |
| **Lactate** (per unit increase) | 2.91 (1.73 – 4.89)* | - | 3.43 (1.10 – 10.7)* |
| **Measures of quality of logistic regression models** |  |  |  |
| • Pseudo-R2 | - | 0.46 | 0.60 |
| • Hosmer and Lemeshow | - | 0.33 | 0.63 |
| • Akaike Information Criterion (AIC) | - | 0.71 | 0.90 |
| • ROC area |  | 0.92 | 0.95 |

\*95% confidence intervals do not cross 1 ( $p < 0.05$ )
T1RF: Type 1 Respiratory Failure, SpO2/eFiO2: pulse oximetry to estimated fraction of inspired oxygen, NLR: neutrophil-lymphocyte ratio, eGFR: estimated glomerular filtration rate, ROC: receiver-operator curve

Results of the pre-defined sensitivity analyses were consistent with the results of the main analysis (see Table 4).

**Table 4.** Sensitivity analyses.

|  | Model 1 | Model 2 |
| --- | --- | --- |
| **COVID – 19 confirmed cases only** | n=126 | n=116 |
| **T1RF (SpO2/eFiO2 ratios)** |  |  |
| None (>316) | 1 (reference) | 1 (reference) |
| Mild (316 – 232) | 1.26 (0.22 – 7.02) | 0.28 (0.02 – 3.37) |
| Moderate (231 – 148) | 5.68 (1.20 – 26.9)* | 4.96 (0.67 – 36.5) |
| Severe (<148) | 62.8 (7.52 – 525)* | 43.1 (1.97 – 942)* |
| **Binary classification of T1RF (SpO2/eFiO2 ratio)** | n=158 | n=155 |
| None/mild (>190) | 1 (reference) | 1 (reference) |
| Moderate/Severe (<=190) | 11.1 (3.48 – 35.6)* | 7.22 (1.75 – 29.9)* |

\*95% confidence intervals do not cross 1 ( $p<0.05$ )
T1RF: Type 1 Respiratory Failure, SpO2/eFiO2: pulse oximetry to estimated fraction of inspired oxygen

## DISCUSSION

In this prospective analysis of patients admitted with suspected COVID-19, a pragmatic classification of T1RF, based on the SpO2/eFiO2 ratio in the ED, was a strong and independent predictor of inhospital mortality. Other predictors of mortality were age, raised lactate levels and reduced renal function as measured by eGFR. Of note, within the limitations of this research, no other vital parameters or blood tests were clearly independently associated with mortality. Results were consistent across different models and sensitivity analyses.

Acute lung injury and an ARDS-like picture are the hallmark of COVID-19 infection and a main cause of mortality.[6] This is confirmed by the sobering statistic of patients presenting to the ED with suspected COVID-19 in our study, with mortality rates of 30% and 68% in patients with moderate and severe T1RF, respectively. Importantly, while the mortality rate in patients with no or mild T1RF (10% and 16%, respectively) are significantly lower, patients in these groups were nevertheless at considerable risk of deterioration and death. While our data did not allow for more detailed analysis of the in-hospital course of illness, it would be of value for future research to investigate if the mortality in the no/mild T1RF patient group is due to a later development of an ARDS-type picture or other pathology, such as venous thrombotic events, cardiac complications or multi-organ failure.[16] This would allow targeted monitoring and appropriate escalation of care, as required.

A number of previous studies have attempted to predict mortality from COVID-19 infection based on factors measured in ED.[5] Importantly, many of these models do not include the presence or severity of T1RF, which in our study has a strong association with mortality. Clinicians in the ED should be cautious in applying the results of models which are ‘blind’ to important clinical features which are readily available to the clinician, such as T1RF. For example, NLR is a frequently cited predictor of mortality in patients with COVID-19.[17, 18] However, in our study, NLR was associated with mortality only on univariate analysis but not once adjusted for the severity of T1RF. While our study was not powered to address this question specifically, we also noticed a similar effect with comorbidities such as diabetes and chronic kidney disease (Appendix 1). This raises the possibility that these are factors which increase the risk of patients presenting to the ED with more severe COVID-19 disease, but which do not necessarily predict further deterioration in themselves. Further, large scale research is required to provide clarity before any of the current predictor models can be applied during clinical care in the ED.(5) We suggest that such further research includes SpO2/eFiO2 ratio or a similar measure of respiratory dysfunction.

Of note, the presence and severity of T1RF as measured by the SpO2/eFiO2 ratio was the only vital parameter in our study which showed an association with mortality. Neither mean arterial blood pressure, heart rate, respiratory rate, nor body temperature were associated with mortality in univariate or multiple logistic regression. Together with either the Glasgow Coma Scale (GCS) or another measure of neurological disability (ACVPU scale), these factors are frequently combined to create early warning scores for deterioration or developing sepsis.[19] In our study, only age, T1RF, eGFR and Lactate levels were associated with in-hospital mortality. Further research is required to examine if the currently used early warning scores are appropriate for inpatients with COVID-19 or should be adapted with an increased focus on the assessment of T1RF.

Finally, as the SpO2/eFiO2 ratio is readily available without requirements for invasive testing or laboratory infrastructure, it might be useful in low resource settings around the world (see Appendix 1 for an example of an SpO2/eFiO2 ratio nomogram).

### Limitations

This was a single centre study of patients admitted to hospital with suspected COVID-19. Due to the limited sample size we did not attempt to create a comprehensive prediction model of mortality or ICU admission but focused on the prognostic value of SpO2/eFiO2 ratio. The calculation of the SpO2/eFiO2 ratio and its application to create T1RF categories contains a number of approximations which likely reduce the accuracy of this measure. The results of this research should be seen as hypothesis generating, rather than providing definite information or directly informing clinical care. The mortality rate in our cohort was relatively high compared to mortality rates of admitted patients reported internationally. This is likely a reflection of common UK practice to discharge patients with suspected COVID-19 without oxygen requirement, if possible, and the relatively advanced age of the cohort in this study.

## Data Availability

Please contact the lead author to discuss data sharing.

## ACKNOWLEDGEMENTS

Dr Neeraja Sritharan and Ms Millie Watkins for their help with data collection.

## COMPETING INTERESTS

The authors have no competing interests to declare.

## Notes

### Competing Interest Statement

The authors have declared no competing interest.

### Funding Statement

No funding was received for this research.

